# Development and Internal Validation of a Large Language Model Pipeline for Multi-Label Classification of Patient Portal Messages

**DOI:** 10.64898/2026.08.14.26360460

**Authors:** Bryan D Steitz, Oluwateniayo O Ogunsan, Jessica Ancker, Brian Carlson, Leslie S Gaynor, Robin T. Higashi, Emily L Morrow, Thomas J Reese, Raymond R. Romano, Sarah Stern, Robert W Turer, S Trent Rosenbloom, Adam Wright

## Abstract

**Objectives:** Characterizing patient portal message content at scale can help target efforts to manage administrative work. We developed and validated a large language model (LLM) pipeline for multi-label classification of messages using an expert-derived topic taxonomy, then characterized topic distribution across a two-year corpus.

**Materials and Methods:** We studied all medical advice request messages sent to ambulatory clinicians at an academic medical center from 2024-2025. We convened an expert panel that derived an 11-category taxonomy through a modified Delphi process. Two annotators labeled 750 randomly selected messages (Cohen kappa 0.80), holding out 500 for evaluation. The pipeline used GPT-4o-mini in a zero-shot prompt. On the held-out set, we measured micro- and macro-averaged precision, recall, and F1, and label stability across runs. We then characterized topic distribution and co-occurrence across the corpus.

**Results:** The pipeline achieved micro- and macro-averaged F1 of 0.89 and 0.86. Labels were identical across runs for 93.6% of messages. Across 2.4 million messages, content concentrated on a few topics. The two most common topics, Problems C Management and Medications C Prescriptions, were present in 67.9% of messages, and the four most common in 93.9%. 51.7% of messages addressed multiple topics.

**Discussion and Conclusion:** The pipeline classified patient message topics accurately and stably across millions of messages. Message content was concentrated within a small number of topics, highlighting opportunities for targeted interventions and enabling more efficient triage, routing, and patient-facing support.

## INTRODUCTION

Asynchronous secure messaging between patients and healthcare professionals via patient portals has become a routine channel for clinical communication. Message volume has grown steadily over the past decade due to health systems expanding patient portal access.[1] Patients have increasingly turned to messaging in place of phone calls and in-person visits.[2, 3] Message volume further increased at the onset of the COVID-19 pandemic and has not returned to pre-pandemic levels.[4] Federal policy has also contributed to this growth. Under the 21^st^ Century Cures Act Information Blocking provision, immediate release of electronic health information has been associated with a near doubling of patient-initiated messages in the hours after patients review their test results.[5, 6]

This increased messaging rate expands access to care but creates administrative burden. For clinicians, managing and responding to patient-initiated messages constitutes a large and mostly uncompensated component of work. This work is a well-acknowledged contributor to clinician exhaustion and burnout.[7] Health systems must manage this tension as care delivery continues to extend into asynchronous channels.[7–10]

Managing messages requires an understanding of the needs and concerns patients express.[11] Prior attempts to characterize message content comprehensively and at scale have leveraged different methods. Unsupervised clustering and topic modeling can identify latent themes[3, 12] but the resulting clusters lack explicit labels and are unstable as message content changes over time. Supervised approaches instead classify messages into investigator-derived categories, and foundational work using these approaches has sorted messages into broad communication types, such as information, medical, logistical, and social.[13–16] However, these broad categories lack the granularity needed for many downstream tasks. More recent studies have achieved strong performance within narrow domains, such as detecting mental health concerns, but these domain-specific models cannot characterize the diversity of content needed to address the broader care needs of patients and the content requested of clinicians.[17–19] Characterizing message content at scale requires an approach that applies consistent, interpretable categories across a full breadth of a general message corpus to enable downstream interventions.

Large language models (LLMs) are trained on vast text corpora to process and generate human-like text across domains. Their application has expanded to clinical tasks such as documenting encounters,[20, 21] summarizing records,[22] and drafting responses to patient messages.[23–25] Applied to patient portal messages, LLMs offer an opportunity to classify content at scale, supporting downstream tasks such as triage, routing, and in basket-related quality improvement. In this study, we developed and evaluated a HIPAA-compliant LLM pipeline for multi-label classification of patient portal messages using an expert-derived taxonomy of message topics. We validated the pipeline against a human gold standard and characterized error patterns. We then applied the pipeline to a corpus of patient-initiated messages spanning two years to describe topic distribution and co-occurrence.

## METHODS

We conducted this study at Vanderbilt University Medical Center (VUMC), a large academic medical center that sees over 3.2 million visits annually in more than 180 ambulatory locations across central Tennessee.[26] Patients at VUMC can review electronic health information, message their clinicians, and perform other administrative functions through My Health at Vanderbilt (MHAV), VUMC’s MyChart-based patient portal (Epic Systems Corp).[27] This study was approved and granted a waiver of consent by VUMC’s Institutional Review Board. We followed the Transparent Reporting of a Multivariable Prediction Model for Individual Prognosis or Diagnosis (TRIPOD-LLM) reporting guideline.[28]

### Study Design and Population

We developed and evaluated an LLM-based pipeline to classify the content of patient portal messages. Our study sample comprised all Epic-indicated patient medical advice request messages sent to an ambulatory clinician at VUMC from January 1, 2024 through December 31, 2025. We included only the message that initiated each thread, sent by patients aged 18 years or older or their authorized proxies. We excluded messages written in a language other than English. The individual message served as our unit of analysis. We extracted all study data from Epic’s Clarity reporting database.

### Development of Classification Taxonomy

To classify message content, we developed a new taxonomy of patient information needs for this study. We aimed to capture core topics of patient need rather than clinical resources required to manage a given message or need specific to a single specialty or population. We derived the taxonomy using a modified Delphi process with a panel of 8 experts, including 3 patient messaging researchers, 4 practicing physicians, and 1 nurse. We seeded the process with the taxonomy of Consumer Health Information Needs[13], from which one patient messaging researcher derived an initial set of core categories representing the most frequent patient message topics. We distributed this set to the remaining panelists electronically and solicited feedback and proposed revisions.

We continued refining the categories across iterative rounds until panelists proposed no further changes. We resolved all discrepancies through discussion until the panel reached unanimous agreement. The final taxonomy comprised 11 categories and is presented in Table 1.

**Table 1.** Taxonomy of Patient Information Needs Expressed via Portal Messaging.

| Topic | Description |
| --- | --- |
| Appointments & Scheduling | Scheduling, rescheduling, or confirming an appointment or visit. |
| Contact Information | Providing or requesting specific contact details, such as phone number, address, or fax number. |
| Forms & Documents | Request to complete or provide forms, letters, paperwork, or other documentation, which may or may not include an attached image or file. |
| Imaging & Tests | Scheduling, availability, or interpretation of a laboratory, imaging, or diagnostic test or result. |
| Insurance & Billing | Providing or requesting details about insurance coverage, prior authorization or billing. |
| Medical Equipment | Requesting details about medical equipment, assistive devices, or implants. |
| Medications & Prescriptions | Requesting details about a specific medication, vaccine, or drug class, including refills, side effects, dosing, and new prescriptions. |
| Personnel & Referrals | Requesting a referral or information about a specific provider or service. |
| Procedures | Scheduling, preparation, or recovery for a surgical or invasive procedure. |
| Problems & Management | Reporting a symptom or request for help managing a new or existing health concern. |
| Other/None of These | Content that fits no other category. |

### Gold Standard

To construct our gold standard, we randomly selected 750 messages from the study cohort and imported them into REDCap[29] for annotation. Two annotators with expertise in clinical informatics and medicine (BDS and OOO) independently labeled each message with one or more topics from the taxonomy. We assessed interrater reliability by computing Cohen kappa for each topic and averaging across the 11 topics, which yielded a mean kappa of 0.80. The two annotators then met to reconcile each discrepancy through discussion and reached consensus on final labels for each message. We randomly partitioned the resulting gold standard into a development set of 250 messages, used to refine the classification prompts, and a held-out evaluation set of 500 messages.

### Natural Language Processing Approach

We developed an automated pipeline to assign multi-label classifications to each message using GPT-4o-mini (version 2024-07-18), accessed through a HIPAA-compliant Azure OpenAI (Microsoft Co.) deployment so that message text could be processed without removing protected health information. We selected GPT-4o-mini after comparing its classification performance with GPT-5.4-mini (version 2026-03-17) on the held-out evaluation set (Supplemental Table 1). GPT-4o-mini achieved marginally higher performance at substantially lower cost (Supplemental Table 2).

Before classification, we stripped each message of non-printable control characters. We then submitted each message to the model within a zero-shot prompt (Supplemental Box 1) that presented the 11 topic definitions and instructed the model to return every applicable label as a structured JSON response. We set the temperature to 0 and top-p to 1.0, configuring the model for greedy, near-deterministic decoding. We iteratively refined this prompt on the development set and report final pipeline performance on the held-out evaluation set.

After establishing pipeline performance on the held-out evaluation set, we applied the validated pipeline to the remainder of the message corpus. We classified all remaining messages in our corpus using the Azure OpenAI Batch API, which processes requests asynchronously, allowing us to label the corpus efficiently at scale. We computed the total classification cost using the published batch API rates as of July 2026: $0.075 per million input tokens and $0.30 per million output tokens.

### Statistical Analysis

We evaluated classification performance on the held-out 500-message evaluation set by comparing the pipeline-assigned labels with the gold-standard labels. To account for run-to-run variation in the modeling pipeline, we classified the evaluation set five times under the production configuration and computed each metric on every run. Across the entire sample, we report exact-match accuracy (the proportion of messages whose complete predicted label set matched the gold-standard set) and micro- and macro-averaged precision, recall, and F1. We computed both averages across the 10 substantive topics. We excluded Other/None of These as it captured residual messages that, after manual review, were deemed acknowledgements or ambiguous messages without a specific need. Micro-averaged metrics pooled true positives, false positives, and false negatives across these topics, and macro-averaged metrics were the unweighted mean of per-topic values. We also summarized precision, recall, and F1 for each topic. Each metric is reported using mean and 95% confidence intervals across the five runs.

We additionally assessed prediction stability across the five runs. We measured stability as the proportion of messages assigned an identical label set in all five runs and report a per-topic disagreement rate describing how often each topic’s assignment changed across runs. To characterize failure modes, we reviewed the evaluation-set messages whose predicted labels disagreed with the gold standard and present paraphrased examples of common false positive and false negative patterns. We constructed an error-linked heatmap to characterize false negatives.

For each message, we paired each false negative topic with any false positive topics predicted on the same message. We attributed a false negative to a substituted topic when a co-occurring false positive was present and to no substitution otherwise. When a message contained multiple false negatives or false positives, we counted each co-occurring pair.

We applied the validated classification pipeline to the full two-year corpus to describe the distribution of message topics. We report the frequency and proportion of each topic across the corpus and the co-occurrence of categories within messages.

## RESULTS

### Classification Performance

Our gold standard evaluation set of 500 messages contained a median of 50 words per message (IQR 28-81) and an average of 1.6 topics per message (SD 0.67). The proportion of messages assigned two or more topics was 49.2%. The most common topics were Medications & Prescriptions (220; 44.0%), Problems & Management (203; 40.6%), and Appointments & Scheduling (98; 19.6%). Overall, the pipeline achieved an exact-match accuracy of 0.738 (95% CI, 0.729-0.746). Across the 10 substantive topics, excluding Other/None of These, micro-averaged precision, recall, and F1 were 0.885 (95% CI, 0.882-0.889), 0.895 (95% CI, 0.890-0.899), and 0.890 (0.887-0.893). Macro-averaged values were 0.874 (95% CI, 0.868-0.880), 0.857 (95% CI, 0.844-0.870), and 0.863 (95% CI, 0.853-0.872). Per-topic performance is reported in Table 2. Across the 10 substantive topics, Medications & Prescriptions yielded the highest F1 score of 0.934, followed by Insurance & Billing (0.933), Appointments & Scheduling (0.917), and Imaging & Tests (0.913).

**Table 2.** Overview of Classification Performance, per Topic. Cross-run disagreement reflects the proportion of messages within a single topic whose assignment was not identical across all runs.

| Topic | Gold-Standard Frequency | Precision (95% CI) | Recall (95% CI) | F1 Score (95% CI) | Cross-Run Disagreement |
| --- | --- | --- | --- | --- | --- |
| Appointments & Scheduling | 98 | 0.958<br>(0.944-0.972) | 0.880<br>(0.874-0.885) | 0.917<br>(0.912-0.922) | 0.014 |
| Contact Information | 48 | 0.883<br>(0.863-0.902) | 0.908<br>(0.894-0.923) | 0.895<br>(0.879-0.912) | 0.008 |
| Forms & Documents | 39 | 0.866<br>(0.851-0.880) | 0.923<br>(0.923-0.923) | 0.893<br>(0.886-0.901) | 0.004 |
| Imaging & Tests | 95 | 0.942<br>(0.924-0.960) | 0.886<br>(0.880-0.892) | 0.913<br>(0.902-0.924) | 0.016 |
| Insurance & Billing | 31 | 0.966<br>(0.966-0.966) | 0.903<br>(0.903-0.903) | 0.933<br>(0.933-0.933) | 0.000 |
| Medical Equipment | 17 | 0.967<br>(0.910-1.000) | 0.741<br>(0.630-0.852) | 0.838<br>(0.746-0.930) | 0.016 |
| Medications & Prescriptions | 220 | 0.945<br>(0.941-0.950) | 0.923<br>(0.923-0.923) | 0.934<br>(0.932-0.936) | 0.010 |
| Personnel & Referrals | 18 | 0.802<br>(0.780-0.825) | 0.856<br>(0.818-0.893) | 0.828<br>(0.808-0.848) | 0.004 |
| Procedures | 24 | 0.609<br>(0.569-0.650) | 0.633<br>(0.610-0.656) | 0.621<br>(0.596-0.645) | 0.012 |
| Problems & Management | 203 | 0.805<br>(0.800-0.811) | 0.913<br>(0.904-0.923) | 0.856<br>(0.851-0.861) | 0.032 |
| Other/None of These | 6 | 1.000<br>(1.000-1.000) | 1.000<br>(1.000-1.000) | 1.000<br>(1.000-1.000) | 0.000 |

Predictions were highly stable across the five runs, with 93.6% of messages receiving an identical set of labels in all runs. The per-topic disagreement rate among these topics was lowest for Insurance & Billing (0.000) and highest for Problems & Management (0.032).

We characterized classification errors across 5 evaluation runs using an error-linked heatmap in Figure 1. Across all topics, the model failed to make any assignment in 54.9% of false negative predictions. This pattern was most pronounced for Problems & Management (77.3%), Imaging & Tests (63.0%), Procedures (56.8%), and Medications & Prescriptions (55.3%). For the remaining six topics, more than half of false negative predictions were linked to a substituted topic. Problems & Management was the most common false positive topic, at a mean of 44.8 false positives per run. In most cases, the model added Problems & Management as an extra label to messages with correct topics already identified. In an average of 16.8 false positives per run, the model applied Problems & Management in place of a topic it missed. Most often, these were labeled in place of Medications & Prescriptions (7.0), Imaging & Tests (4.0), and Procedures (3.0). Procedures showed the highest false positive rate relative to its prevalence at a mean of 9.8 false positives per run against 24 true instances. We provide example messages with false positive and false negative predictions in Supplemental Table 3.

**Figure 1.**
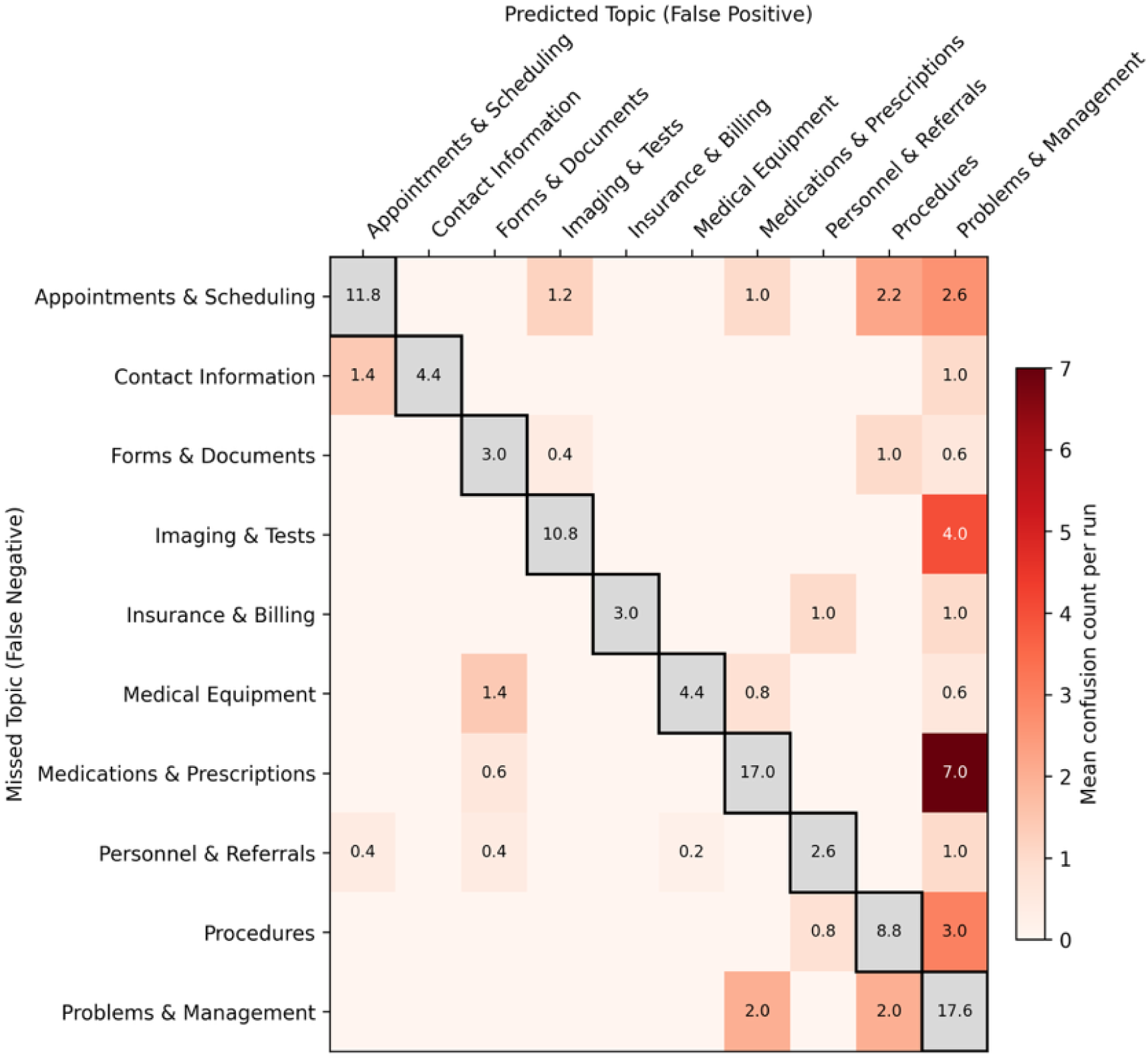
Heatmap of Incorrect Predictions. Values are averaged across 5 runs. Diagonal cells (grey) show the mean false negative count for that topic and are not on the same color scale as the off-diagonal cells.

### Corpus Topic Distribution

The 2024-2025 message corpus contained 2,416,394 messages which we classified using our pipeline. Among these, 176 messages could not be classified due to the LLM content filter. This left 2,416,218 messages sent by 379,768 patients to 5,361 clinicians. These messages contained 3,881,537 topics with an average (SD) of 1.6 (0.66) topics per message (Figure 2). The most common message topics were Problems & Management (1,026,978; 42.5%), Medications & Prescriptions (898,255; 37.2%), and Imaging & Tests (509,770; 21.1%). Message content was concentrated within a small number of topics. The two most frequent topics, Problems C Management and Medications C Prescriptions, were present in 67.9% of messages. The three most frequent topics were present in 82.7% of messages, and the four most frequent topics in 93.9% of messages. Nearly half the messages contained a single topic (1,166,113; 48.3%), and only 196,582 (8.1%) messages contained three or more topics (Figure 3). Among single-topic messages, the most common were Problems & Management (311,681; 26.7%), Medications & Prescriptions (306,052; 26.2%), Imaging & Tests (128,669; 11.0%) and Appointments & Scheduling (126,859; 10.9%). Among multi-topic messages, the most frequent pairings were Problems C Management with Medications C Prescriptions (283,943; 22.7%), with Imaging & Tests (124,686; 10.0%), and with Appointments & Scheduling (66,965; 5.4%). Running this pipeline across all messages incurred a total cost of $123.50.

**Figure 2.**
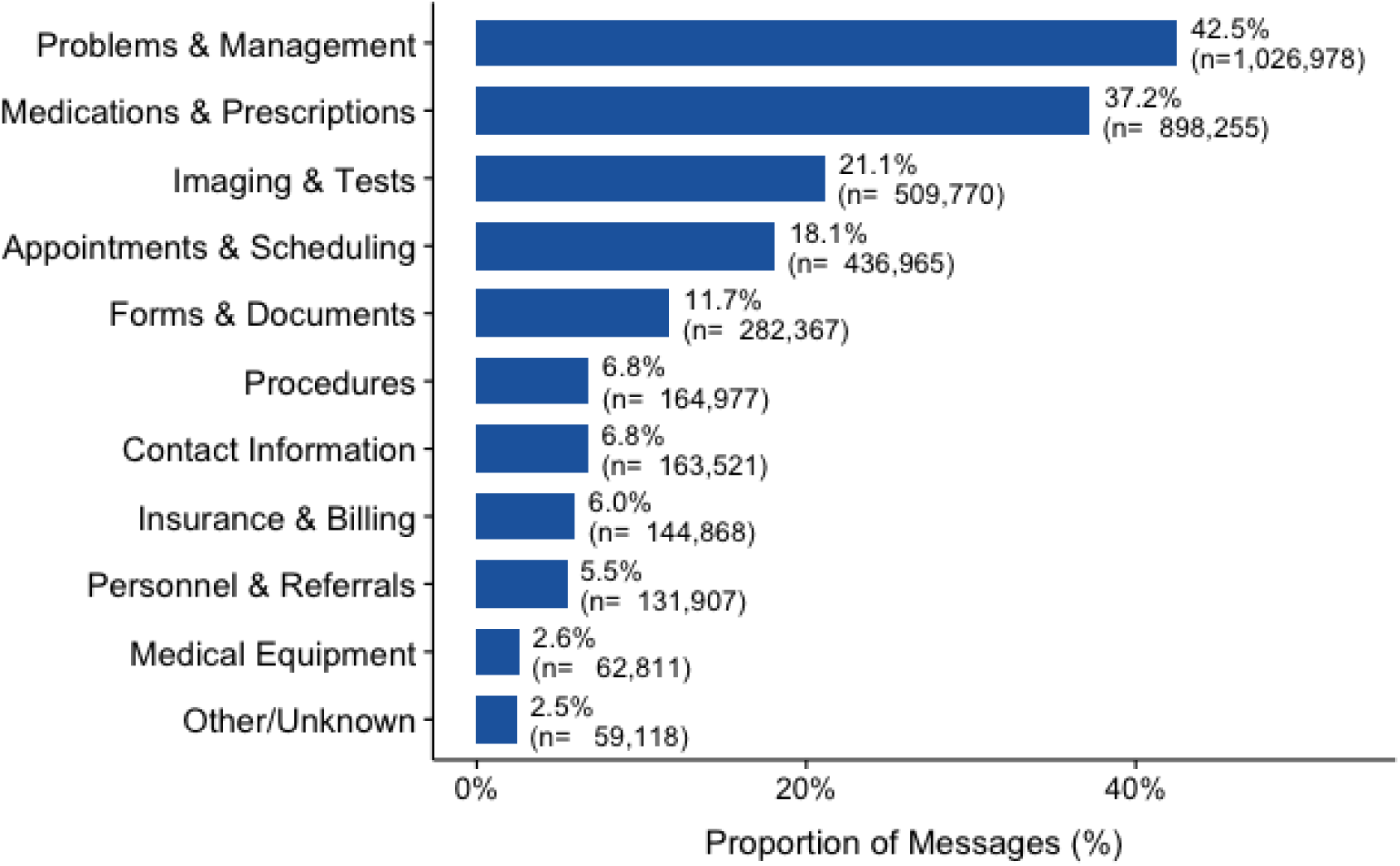
Distribution of Message Topics within Patient-Initiated Messages. Percentages represent the proportion of messages containing the respective topic.

**Figure 3.**
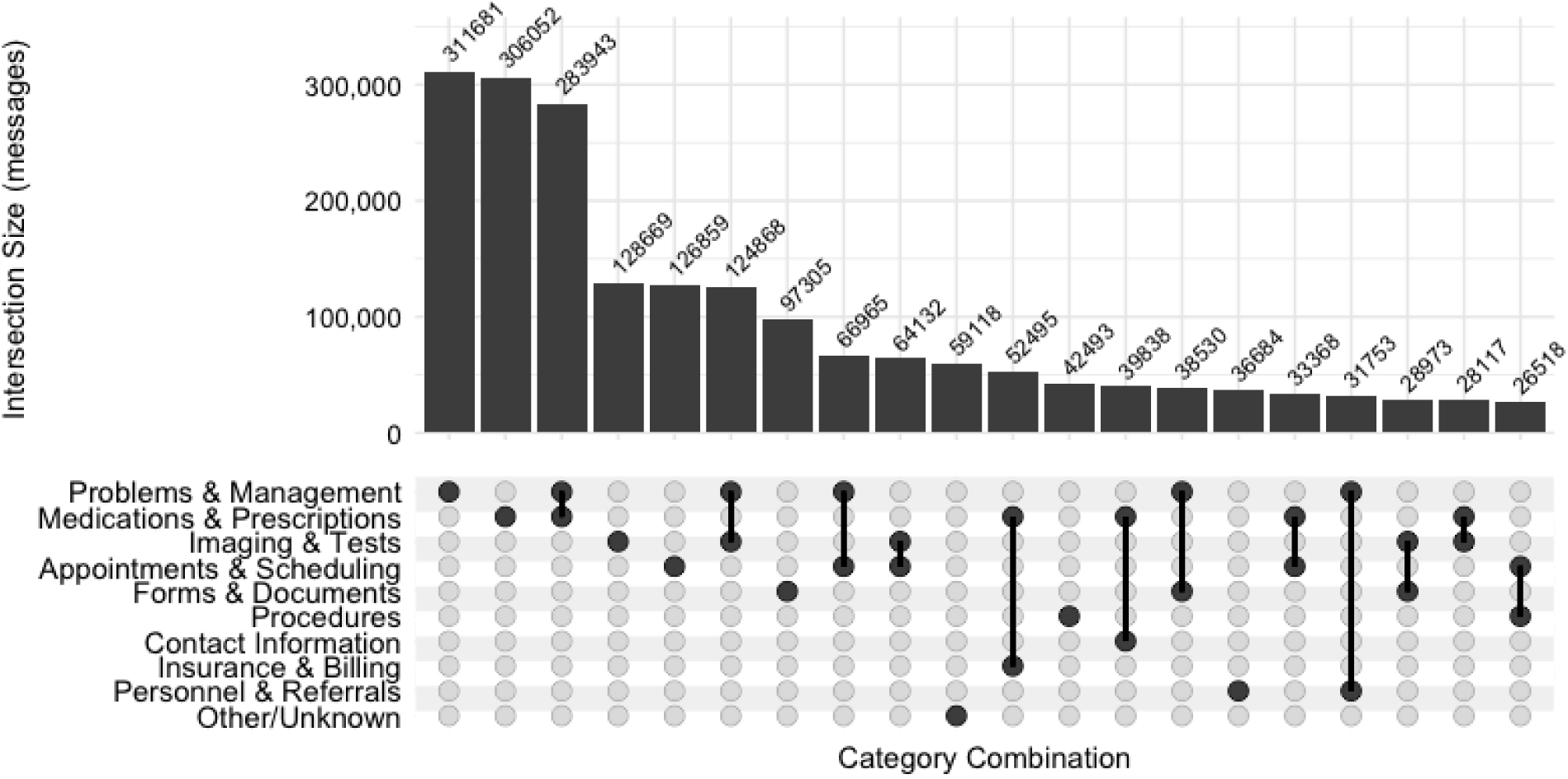
UpSet Plot of Top 20 Topic Co-Occurrence. Each row in the dot plot represents a topic classification and connecting solid dots represent each topic part of an intersecting set. The bar graph depicts the number of messages in each intersection.

## DISCUSSION

In this study, we developed and validated an LLM pipeline that classifies the content of patient portal messages into 11 expert-derived topics. We applied the validated pipeline to a corpus of over 2.4 million messages spanning 2 years from a large academic medical center. The pipeline classified messages with strong agreement against a human gold standard of 500 messages, achieving a micro-averaged F1 of 0.89 and macro-averaged F1 of 0.86. Across the full corpus, message content was highly concentrated on a few topics. We observed that Problems C Management and Medications C Prescriptions were the most frequent categories. Together, they were present in 67.9% of messages; 93.9% of messages contained at least one of the four most common topics. Characterizing at scale the topics that drive message volume is an integral first step toward helping health systems manage message-related work.

This work extends prior studies which have characterized messages by communication type.[13–15] Prior work categorized messages as information (requiring clinical knowledge), medical (requests for delivery of care), logistical (pragmatic, non-clinical matters), or social (interpersonal communication).[13] These communication types indicate the type of work a message requires, but not the specific need that a patient raises.[8] Our taxonomy instead classifies messages by topic, a complementary framing that aligns with patient needs rather than clinical work. A single topic in our taxonomy may span multiple communication types. Messages about Medications & Prescriptions may request clinical information, ask for a refill, or report an issue with the pharmacy. Conversely, our topics resolve distinctions that these broader communication types collapse, separating Imaging C Tests, Appointments C Scheduling, and Personnel & Referrals into individual topics. Current approaches to triage and routing commonly rely on manually defined rules and broad patient-selected categories, such as a medical or prescription question, that are coarse and inconsistently applied. Furthermore, these messages are often broadly routed to staff for initial review before those requiring a clinician are forwarded. By naming the subject of each message, our categories align more closely with current triage and routing practices, which direct messages by topic and responsible team member rather than by communicative function.[30] Characterizing both the topic of a message and the work it requires may support more precise redirection as systems grow in their ability to handle greater complexity. Combining topics with measures of clinical effort to redistribute message workload is an important direction for future work.

Our taxonomy is intended to capture core message topics rather than provide an exhaustive inventory of patient information needs, many of which are specific to populations or clinical specialties. Prior work has applied clustering and topic modeling to characterize message content.[3, 12, 18, 31] However, applied to a broad, heterogeneous corpus, these methods commonly recover unstable topics across runs and often produce clusters without explicit or readily interpretable labels.[32, 33] Applied within a single topic where content is more homogeneous, clustering can instead resolve coherent subthemes and reveal micro-level trends.[34, 35] Within Medications & Prescriptions, for example, clustering could separate refill requests from reports of side effects or questions about medication administration. Combining the stable, interpretable, expert-derived taxonomy with within-topic clustering is an important extension of this work and a direction of future study.

Growing patient message volume is a known contributor to professional exhaustion and burnout among physicians and nurses who share the work of triaging and responding to messages.[7, 36] However, interventions that aim to manage message-related work often neglect message content. Messages are also increasingly complex, often raising several needs at once. In our corpus, messages carried an average of 1.6 topics, and over half raised two or more topics. Efforts to manage message burden have relied on routing and staffing models that intend to redistribute message work from providers to clinical staff using static routing rules or a centralized pool-based approach.[30, 37] Without message content to guide routing, a message can reach a team member unable to resolve it, who must then forward it to someone else or return to the patient for clarification – adding to the message volume. Recognizing message topics at the point of triage could allocate this work more precisely, directing each message to the staff member or appropriate workflow best suited to its content. Further, pairing topics with the classification of the anticipated work a message requires could direct clinical requests to the staff best suited to handle them and route administrative requests to automated work queues. Appointments & Scheduling, for example, accounted for 126,859 (10.9%) single-topic messages, indicating that patients raise scheduling needs as advice requests even where structured scheduling workflows exist. Topics that are reliably non-clinical, including Insurance C Billing, Forms C Documents, and Contact Information could similarly be directed to administrative staff or digital agents without clinician review.

Understanding message content can also identify common questions that may be addressed outside of messaging workflows. Immediate release of test results under the information blocking provisions of the 21st Century Cures Act was associated with a near doubling in patient-initiated messages in the hours following result review.[6] Interventions to manage this message volume have largely focused on managing result release workflows rather than unmet information needs,[5] despite research showing that nearly 60% of patients seek additional information to understand their results.[38] We found that test-related messages accounted for 21.1% of messages in our corpus. Classifying this content at scale identifies where result-related questions concentrate. It can allow health systems to characterize the questions patients raise about their results and to surface targeted education, addressing common concerns proactively.[39] Message burden is not one-sided. As volume outpaces the capacity to reply, patients also experience delayed or incomplete responses. Interventions that manage messages more efficiently serve patients as well as the care team.

This study has several limitations. First, it was conducted at a single academic medical center with messages in English only. Message content and topic distribution may differ across institutions, patient populations, and portal configurations. Second, we classified only medical advice request messages. These messages are the primary source of free-text patient concerns, whereas requests such as medication refills or scheduling are often handled through structured workflows. Third, we analyzed only the messages that initiated each thread. Each thread is intended as a unit of discussion around a single topic or need. However, some patients may surface additional needs in subsequent messages within a thread, and these needs are not reflected in our message corpus. Fourth, we evaluated the pipeline against a gold standard of 500 held-out messages. Low-prevalence categories contained relatively few instances, which reduced the precision of their performance estimates. Finally, we classified messages using GPT-4o-mini to balance language model performance with sufficiently low cost such that the pipeline could be applied at scale across an organization to support ongoing operational improvement. Recent comparisons have found that modern LLMs improve classification of patient texts compared with existing NLP-based approaches such as BERT and ClinicalBERT.[40, 41] We compared GPT-4o-mini against a single newer model, and other models may yield different performance.

## CONCLUSIONS

We developed and validated an LLM pipeline that classifies the topics contained within patient portal messages and applied it to two years’ worth of messages from a large academic medical center. The pipeline achieved strong agreement with a human-annotated gold standard, with a micro-averaged F1 of 0.89, and produced stable classifications across repeated runs. Applied to more than 2.4 million messages, the pipeline showed that message content was highly concentrated, with two topics present in 67.9% of messages and four topics in 93.9%. A topic-level view enables data-driven triage, routing, and patient-facing interventions, organized by content.

## CONTRIBUTOR STATEMENT

BDS, AW, STR, and JSA conceived the study. BDS extracted the analytic dataset, developed the NLP pipeline, and drafted the initial manuscript. BDS, OOO, LSG, ELM, and RRR developed annotation procedures and performed initial validation of the message annotation pipeline. BDS and OOO annotated the full gold standard dataset. BC, RTH, TJR, SS, and RWT provided guidance on model development, statistical analysis, and clinical interpretation. All authors revised the draft and approved the submitted manuscript.

## DATA AVAILABILITY STATEMENT

Data used in this research may contain protected health information and cannot be shared publicly. The authors will share representative, deidentified, sample messages upon request.

