## Supplemental Materials for "Development and Internal Validation of a Large Language Model Pipeline for Multi-Label Classification of Patient Portal Messages"

**Supplemental Box 1.** Message Classification Prompt
**Supplemental Table 1.** GPT 5.4-mini Classification Performance.
**Supplemental Table 2.** Comparison of Classification Performance, by Category, Across Models.
**Supplemental Table 3.** Sample of Incorrect Predictions

**Supplemental Box 1.** Message Classification Prompt

Classify a patient portal message using one or more of the labels below. Assign a label only when the message is substantively about that topic. Do not add a label because a topic is mentioned as context or background for a different request. Most messages have one or two labels.

APT - scheduling, rescheduling, or confirming routine appointments, visits, or consultations. If scheduling involves a named procedure or surgery, use PRC instead.
CNT - phone numbers, addresses, or fax numbers explicitly provided or requested. Requests to call, mail, or fax something without exchanging actual contact details are not CNT.
DOC - requests or information about forms, paperwork, letters, images, or documentation. Messages indicating an attached image should be classified with this label
TST - assign whenever labs, imaging (CT, MRI, X-ray, ultrasound, mammogram), biopsy, pathology report, vitals, or diagnostic tests are mentioned - scheduling, results, interpretation, or a reported value - regardless of whether this is the message's primary topic.
INS - insurance details, billing, or insurance record updates.
EQP - medical equipment, monitors, assistive devices, or implants.
MED - assign whenever a specific medication, drug class, or vaccine is named or described refills, side effects, dosing, new Rx, whether to continue — regardless of whether medication is the message's primary topic.
REF - an explicit request for a referral or for information about a specific provider.
PRC - scheduling, preparation, or recovery for a specific named surgical or invasive procedure (surgery, biopsy, endoscopy, dental procedure). Applies when the procedure is the subject of the request, such as scheduling it, preparing for it, or managing recovery from it. A procedure or infusion mentioned as context for a medication request or symptom question does not qualify. Diagnostic imaging (CT, MRI, X-ray, ultrasound, mammogram) is TST, not PRC.
PRB - the patient describes a symptom they are experiencing or asks how to manage a health situation. Do NOT assign PRB when a condition is mentioned only as context for a scheduling request, refill, administrative task, or referral — even if the condition is clinically significant.
OTH - none of the above. Use alone, never with another label.

Input is a JSON object mapping ids to message text. Classify each message independently. Return only a JSON object with one entry per id, no other text:
{"<id>": {"d": ["LABEL", ...]}}

**Supplemental Table 1.** GPT 5.4-mini Classification Performance. 95% confidence intervals were obtained by performing 5 classification iterations. Across all categories, macro-averaged F1 was 0.82 (0.82-0.83) and micro-averaged F1 was 0.87 (0.86-0.87).

| Topic | Frequency | Precision (95% CI) | Recall (95% CI) | F1 (95% CI) | Cross-Run Disagreement |
|---|---|---|---|---|---|
| Appointments & Scheduling | 98 | 0.883 (0.868-0.897) | 0.767 (0.747-0.788) | 0.821 (0.804-0.837) | 0.024 |
| Contact Information | 48 | 0.744 (0.737-0.750) | 0.979 (0.979-0.979) | 0.845 (0.841-0.850) | 0.004 |
| Forms & Documents | 39 | 0.778 (0.765-0.791) | 0.790 (0.763-0.816) | 0.784 (0.766-0.801) | 0.006 |
| Imaging & Tests | 95 | 0.769 (0.763-0.776) | 0.989 (0.989-0.989) | 0.866 (0.861-0.870) | 0.006 |
| Insurance & Billing | 31 | 0.909 (0.907-0.910) | 0.961 (0.943-0.979) | 0.934 (0.925-0.943) | 0.002 |
| Medical Equipment | 17 | 0.781 (0.722-0.840) | 0.706 (0.706-0.706) | 0.741 (0.715-0.767) | 0.004 |
| Medications & Prescriptions | 220 | 0.863 (0.855-0.871) | 0.970 (0.967-0.973) | 0.913 (0.909-0.917) | 0.024 |
| Personnel & Referrals | 18 | 0.635 (0.609-0.662) | 0.889 (0.889-0.889) | 0.741 (0.723-0.759) | 0.006 |
| Procedures | 24 | 0.633 (0.599-0.668) | 0.817 (0.788-0.845) | 0.713 (0.692-0.734) | 0.014 |
| Problems & Management | 203 | 0.873 (0.869-0.877) | 0.894 (0.884-0.903) | 0.883 (0.879-0.887) | 0.040 |

**Supplemental Table 2.** Comparison of Classification Performance, by Category, Across Models.

| Topic | Frequency | GPT 5.4-mini F1 (95% CI) | GPT 4o-mini F1 (95% CI) | Change in F1 (4o-mini minus 5.4-mini) |
|---|---|---|---|---|
| Appointments & Scheduling | 98 | 0.821 (0.804-0.837) | 0.917 (0.912-0.922) | 0.096 |
| Contact Information | 48 | 0.845 (0.841-0.850) | 0.895 (0.879-0.912) | 0.05 |
| Forms & Documents | 39 | 0.784 (0.766-0.801) | 0.893 (0.886-0.901) | 0.109 |
| Imaging & Tests | 95 | 0.866 (0.861-0.870) | 0.913 (0.902-0.924) | 0.047 |
| Insurance & Billing | 31 | 0.934 (0.925-0.943) | 0.933 (0.933-0.933) | -0.001 |
| Medical Equipment | 17 | 0.741 (0.715-0.767) | 0.838 (0.746-0.930) | 0.097 |
| Medications & Prescriptions | 220 | 0.913 (0.909-0.917) | 0.934 (0.932-0.936) | 0.021 |
| Personnel & Referrals | 18 | 0.741 (0.723-0.759) | 0.828 (0.808-0.848) | 0.087 |
| Procedures | 24 | 0.713 (0.692-0.734) | 0.621 (0.596-0.645) | -0.092 |
| Problems & Management | 203 | 0.883 (0.879-0.887) | 0.856 (0.851-0.861) | -0.027 |

**Supplemental Table 3.** Sample of Incorrect Predictions

| Topic | Error Type | Message Excerpt | Gold Label | Model Prediction |
|---|---|---|---|---|
| Appointments & Scheduling | False Positive | I was going to call and schedule the scan you want me to get, but there isn't an order in my chart for it. Can you please put an order in for it? | Imaging & Tests | Imaging & Tests; Appointments & Scheduling |
| Appointments & Scheduling | False Negative | I'm in town today, Wednesday and Thursday. I am not doing well. Can you find time to see me? | Appointments & Scheduling; Problems & Management | Problems & Management |
| Contact Information | False Positive | It has been about 3 days I don't believe I have a fever the nausea is not real bad right now but with the diarrhea I probably went 15 times yesterday. Please refill my Imodium to Walgreens on [street] | Medications & Prescriptions; Problems & Management | Contact Information; Medications & Prescriptions; Problems & Management |
| Contact Information | False Negative | I called Navitus to get pricing information, and I definitely want to go with the Freestyle Libre 2 (instead of the Dexcom G6). She said that it requires a Pre Authorization Form from you before the pharmacy will fill it. They said you would have to all to request the PAF at 866-333-2757. My member ID is 00204201. Let me know if you have any questions or need further information from me. Thank you! | Contact Information; Forms & Documents; Medical Equipment; Insurance & Billing | Forms & Documents; Medical Equipment; Insurance & Billing |
| Forms & Documents | False Positive | Can you please share my bone density results with [doctor]? | Imaging & Tests | Forms & Documents; Imaging & Tests |
| Forms & Documents | False Negative | I had an appointment scheduled for a prosthesis for my left breast. The nurse assisting me stated she could do the fitting due to swelling upon on my chest, which she stated appeared to be fluid. I called the [clinic] and I have appointments scheduled on [date]. I am asking for an extension of medical statement because, I know I will not be able to return to work on [date]]. I am extremely concerned because I will not have my prosthesis and I still have swelling with soreness. | Appointments & Scheduling; Forms & Documents Problems & Management | Imaging & Tests; Problems & Management; Procedures |
| Imaging & Tests | False Positive | My back and hip issues worsened and I received treatment from [clinic]. After X-rays and an MRI the diagnosis is arthritis in my lower spine that impinges nerves causing the back and hip pain. I've received a prescription for an anti-inflammatory, Meloxicam, and wanted to make sure it is not an issue with my blood pressure, statin and low dose aspirin. | Medications & Prescriptions; Problems & Management | Imaging & Tests; Medications & Prescriptions; Problems & Management |
| Imaging & Tests | False Negative | Good morning. Are you going to inform [clinic] about my covid test or do I have to? | Imaging & Tests | Forms & Documents |
| Insurance & Billing | False Positive | When I called to get refill on the following meds they said they need updated prescriptions for refills. Thank you for sending these updates to Champva meds by mail. | Medications & Prescriptions | Insurance & Billing; Medications & Prescriptions |
| Insurance & Billing | False Negative | Please let me know what other dermatologist takes my insurance for a spot on my right hand that is really bad. I went to [clinic] and would rather not be seen there again. | Insurance & Billing; Personnel & Referrals | Personnel & Referrals |
| Medical Equipment | False Negative | Results from interrogation of my defibrillator on [date] indicates battery life only good for approximately 8 months. Should I be concerned, or is there a game plan to replace the battery? | Medical Equipment | Problems & Management |

| | | | | |
|---|---|---|---|---|
| Medical Equipment | False Negative | I was wearing a heart monitor for 2 weeks. I had to stop mid study bc the adhesive strip was severely irritating my skin. I took the strip off Tuesday morning. My skin is inflamed, burns and itches. | Medical Equipment; Problems & Management | Forms & Documents; Problems & Management |
| Medications & Prescriptions | False Positive | I need an immunization certificate filled out for school. What would be the best way to get that done | Forms & Documents | Forms & Documents; Medications & Prescriptions |
| Medications & Prescriptions | False Negative | Is there any cream or ointment I can put on my rash that will help it from being so raw? | Medications & Prescriptions | Problems & Management |
| Personnel & Referrals | False Positive | I have received notice that memantine will not be covered by Medicare as it is being prescribed off label. Can you refer this to prior authorization team? | Insurance & Billing; Medications & Prescriptions | Medications & Prescriptions; Personnel & Referrals |
| Personnel & Referrals | False Negative | In May at my eye appointment with [doctor] at [clinic], he discovered a cataract in my right eye. […] [doctor] referred me to [doctor] who I saw last week. He confirmed the cataract in my right eye and scheduled surgery for [date]. I am reaching out to make you aware of the surgery and to make sure you feel good about this. | Personnel & Referrals Procedures | Procedures |
| Procedures | False Positive | I saved the kidney stone from [date] and will bring it next Monday. Is that the correct procedure? | Procedures | Problems & Management |
| Procedures | False Negative | You said last spring that c19 would spike again in the fall and you were right. Surgery went well, just had more labs drawn with my infusion. Stay well! | Imaging & Tests; Procedures | Imaging & Tests; Problems & Management |
| Problems & Management | False Positive | Can you get me an update on the Dexcom system? I am finding I have to do 40 units of humalog to get my sugar to come down. | Medical Equipment; Medications & Prescriptions | Medical Equipment; Medications & Prescriptions; Problems & Management |
| Problems & Management | False Negative | We are in Nashville for my appointment tomorrow and I realized that I forgot my medication. Can I miss a day or should I drive back home to get it. | Medications & Prescriptions | Medications & Prescriptions; Problems & Management |
